# Scanner-based real-time 3D brain+body slice-to-volume reconstruction for T2-weighted 0.55T low field fetal MRI

**DOI:** 10.1101/2024.04.22.24306177

**Authors:** Alena U. Uus, Sara Neves Silva, Jordina Aviles Verdera, Kelly Payette, Megan Hall, Kathleen Colford, Aysha Luis, Helena S. Sousa, Zihan Ning, Thomas Roberts, Sarah McElroy, Maria Deprez, Joseph V. Hajnal, Mary A. Rutherford, Lisa Story, Jana Hutter

## Abstract

**Purpose:** Integrating the SVRTK methods within the Gadgetron framework enables automated 3D fetal brain and body reconstruction in the low-field 0.55T MRI scanner within the duration of the scan.

**Methods:** A deep-learning based, integrated, robust, and deployable workflow from several motion-corrupted individual T2-weighted single-shot Turbo Spin Echo stacks to produce super-resolved 3D reconstructed fetal brain and body is enabled by combining automated deformable and rigid Slice-to-Volume (D/SVR) reconstruction adapted for low field MRI with a real-time scanner-based Gadgetron workflow. Qualitative evaluation of the pipeline in terms of image quality and efficiency is performed in 12 prospectively acquired fetal datasets from the 22-40 weeks gestational age range.

**Results:** The reconstructions were available on average 6:42±3:13 minutes after the acquisition of the final stack and could be assessed and archived on the scanner console during the ongoing fetal MRI scan. The output image data quality was rated as good to acceptable for interpretation. The additional retrospective testing of the pipeline on 83 0.55T datasets demonstrated stable reconstruction quality for low-field MRI.

**Conclusion:** The proposed pipeline allows scanner-based prospective motion correction for low-field fetal MRI. The main novel components of this work are the compilation of automated fetal and body D/SVR methods into one combined pipeline, the first application of 3D reconstruction methods to 0.55T T2-weighted data, and the online integration into the scanner environment.

## 1 INTRODUCTION

Fetal MRI is an increasingly used adjunct to ultrasound for improved diagnostic accuracy in certain conditions ^1^ as well as providing a detailed characterisation of normal and abnormal fetal development ^2,3^.

Fast acquisition protocols such as T2-weighted (T2w) Half-Fourier Acquisition single-shot Turbo Spin Echo (ssTSE) provide high in-plane 2D image quality ^4^. However, unpredictable fetal motion remains the main limiting factor in both 2D in-plane artifacts and information content related to the loss of 3D structural continuity. Currently, retrospective motion correction performed in the image domain ^5^ has proven to be the most effective solution for structural fetal MRI. Yet, these methods are still primarily in the research stage. Wider integration into diagnostic practice would require a comprehensive formal evaluation of added clinical value as well as optimisation of approaches for deployment directly into clinical settings.

### 1.1 Image-domain 3D reconstruction for fetal MRI

Image-domain motion correction for fetal MRI is based on slice-to-volume registration (SVR) and super-resolution for the reconstruction of high-resolution 3D isotropic images from multiple motion-corrupted stacks acquired under different orientations. Originally proposed over a decade ago for the fetal brain ^6^, these methods evolved into fast deep-learning automated pipelines for the whole fetus.

In addition to the SVR step, the main components of these motion correction methods include super-resolution reconstruction ^7^, intensity matching, and regularisation and rejection of outliers ^8^. While brain reconstruction relies only on rigid registration, motion correction for the fetal body (trunk) requires deformable slice-to-volume registration (DSVR) due to the presence of non-rigid motion ^9,10^. Several works also proposed solutions for automated slice and stack quality assessment and its impact on reconstruction quality ^11,12,13,14^.

More recent research directions primarily focus on using deep learning for the automation of masking, pose estimation, registration, and super-resolution reconstruction. Automation of masking of the fetal brain ^15^ and body ^13^ resulted in fully automatic reconstruction pipelines without manual input. Various pose estimation solutions ^16,17,18,13^ allow both reorientation of the fetal brain and body to the standard space and correction of extreme rotation motion for improved reconstruction quality. Deep learning-based registration and super-resolution reconstruction reportedly result in improved image quality and faster performance ^19,10,20^.

### 1.2 Integration into scanner environment

Integrating image reconstruction and analysis workflows into clinical processes is crucial for translation and to be able to test such image processing pipelines in broader patient populations. However, challenges that hinder wider clinical adoption remain: many published algorithms do not include source code or rely on proprietary accessory code (e.g., vendor-provided), hindering reproducibility. Furthermore, integration into the clinical workflow requires modification of the normal vendor-provided reconstruction pipelines and often requires high-performance computing (GPU) resources.

Several fetal MRI research works reported scanner-based solutions for real-time brain tracking ^21^ and automated detection and re-acquisition of low-quality slices ^22^. Both solutions implemented interaction between the scanner and external GPU-accelerated research reconstruction workstations.

Existing SVR reconstruction methods are performed offline after the fetal scan is completed (i.e., using downloaded NIfTI or DICOM files) in research facilities and there have been no proposed solutions integrated directly into the scanner environment. This is one of the factors limiting the application of 3D fetal reconstruction for clinical reporting.

Gadgetron has been developed as an open-source, modular software platform for sending and retrieving data to/from the scanner reconstruction environment/an external server, respectively. This provides a flexible environment for implementing and executing various image reconstruction and processing algorithms while exploiting the computing power of external servers. The Gadgetron framework has successfully facilitated the integration of several image reconstruction and analysis pipelines into scanner environments, allowing cloud-based reconstruction of free-breathing motion-corrected cine images, quantification of myocardial perfusion, and in-line segmentation of cardiac cavities, among many others ^23,24,25,26^.

### 1.3 Fetal MRI at low field strength

Traditionally performed at 1.5T and 3T, low-field fetal MRI at 0.55T is a re-emerging ^27^ modality for fetal imaging. The increased homogeneity of the magnetic field decreases magnetic susceptibility artifacts frequently found between the fetal brain and the maternal bowel and thereby also the need for specialist shimming tools as commonly used on higher field ^28^. Furthermore, emerging commercial 0.55T scanners offer the benefit of a larger bore size - enabled by the reduced field strength requirement - significantly widening access to fetal MRI. However, the drawback of reduced signal-to-noise ratio has to be taken into account when developing low-field specific techniques.

Recent works ^29,30^ demonstrated the general feasibility of 0.55T MRI for brain and body imaging as well as several examples of fetal brain SVR reconstruction and functional fetal brain and body data reconstruction ^31,32^. Yet, there has been no extensive evaluation of D/SVR performance (or dedicated solutions) to confirm the general feasibility of using 3D reconstruction at 0.55T on a large scale.

### 1.4 Contributions

This work integrates a 3D brain+body D/SVR reconstruction pipeline for T2w structural low field 0.55T fetal MRI integrated directly into the scanner environment and makes the final 3D reconstructions available during the ongoing fetal scan. It combines our previous works on automated 3D SVR reconstruction ^13,32^ with a real-time integrated scanner workflow ^21,33^. In the context of this work, real-time integration refers to the triggering of the SVR reconstruction process immediately upon the acquisition of the last T2w stack, with retrieval of the resulting volume in the duration of the scan.

The reconstruction pipeline is based on the classical methods from SVRTK^*^ toolbox with deep learning automation based on MONAI tools ^34^. It is deployed into the scanner environment via Gadgetron^†^ with the processing being launched directly from specific sequences during the scanning session. The resulting 3D reconstructed files are sent back to the scanner.

The feasibility of the proposed automated D/SVR reconstruction pipeline is evaluated (both quantitatively and qualitatively) on 83 retrospective 0.55T T2w datasets. Evaluation of the scanner deployment is based on real-time (*in utero*) testing on 12 prospective cases in terms of general operability, time, quality of the results, and operator experience.

## 2 METHODS

In this automated workflow, via Gadgetron, the ssTSE images are exported to an external GPU-accelerated server (“Gadgetron server”) in real-time and converted to NIfTI format. Upon the collection of all ssTSE stacks, a 5-second dummy sequence is run with the sole purpose of launching the SVR docker container on the Gadgetron server. Once the D/SVR results are available, short dummy sequences are added to the exam card to pull the resulting volumes to the MRI scanner and store them in the medical image database. This workflow not only allows D/SVR to be run for all patients scanned in the low-field scanner automatically and immediately, with results stored in the database alongside the acquired images, but it also reduces the workload on radiographers and researchers who run the processing pipeline offline post-scan. An overview of these steps is given in Fig. 1 and all individual steps will be outlined in detail below.

**FIGURE 1.**
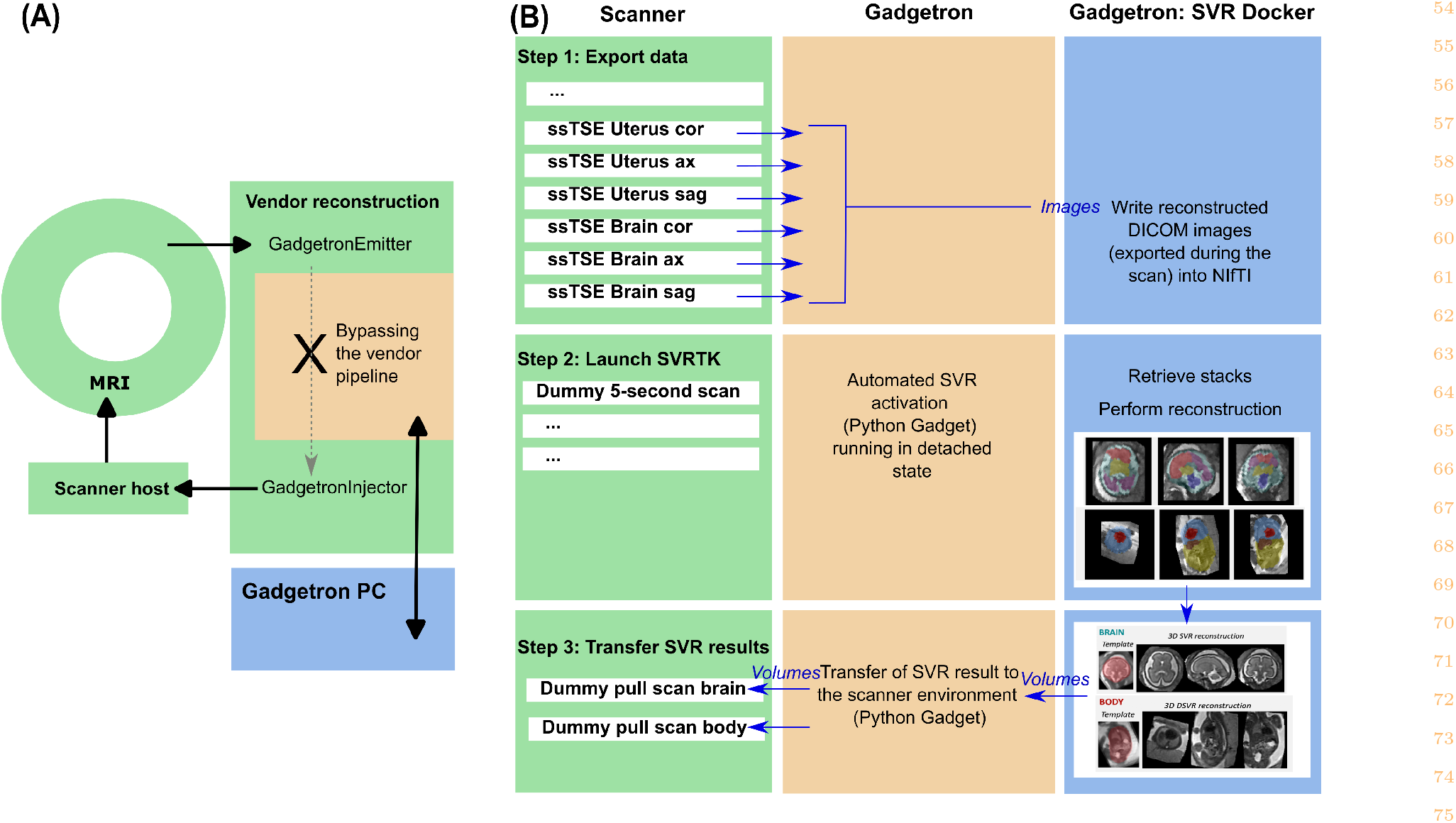
Proposed pipeline for the integration of 3D brain and body D/SVR reconstruction into scanner environment via Gadgetron. (A) Infrastructure setup. (B) Three-step process and flow of images and volumes between the scanner, during the acquisition of the single-shot Turbo Spin Echo scans in coronal, axial and sagittal orientations, the Gadgetron server, and the SVR container within the Gadgetron server.

### 2.1 Datasets, Acquisition

The fetal MRI data used in this study were acquired at St.Thomas’ Hospital, London as part of the ethically approved MEERKAT [REC: 21/LO/0742], MiBirth [REC: 23/LO/0685] and NANO [REC: 22/YH/0210] studies. All experiments were performed in accordance with relevant guidelines and regulations. Informed written consent was obtained from all participants.

The acquisitions were performed on a contemporary clinical 0.55T scanner (MAGNETOM Free.Max, Siemens Healthcare, Erlangen, Germany) with 6-element flexible coil (BioMatrix Contour Coil, Siemens Healthcare, Erlangen, Germany) and a 9-element spine coil built into the patient table. The structural T2w stacks were acquired using a dedicated ssTSE sequence optimised for fetal imaging at 0.55T (low-field) ^29^ with TR = 1460–2500 ms, TE = 105–106 ms, 1.48 mm in-plane resolution, 4.5 mm slice thickness. Each dataset includes 6-12 stacks covering the whole uterus, brain, and trunk regions using standard radiological orientations. The stacks were acquired in consecutive order without time gaps or changes in the maternal position.

The 0.55T datasets used in this work were acquired between 2022 and 2024 and cover 20-40 weeks gestational age (GA) range (Fig. 2) including:

**FIGURE 2.**
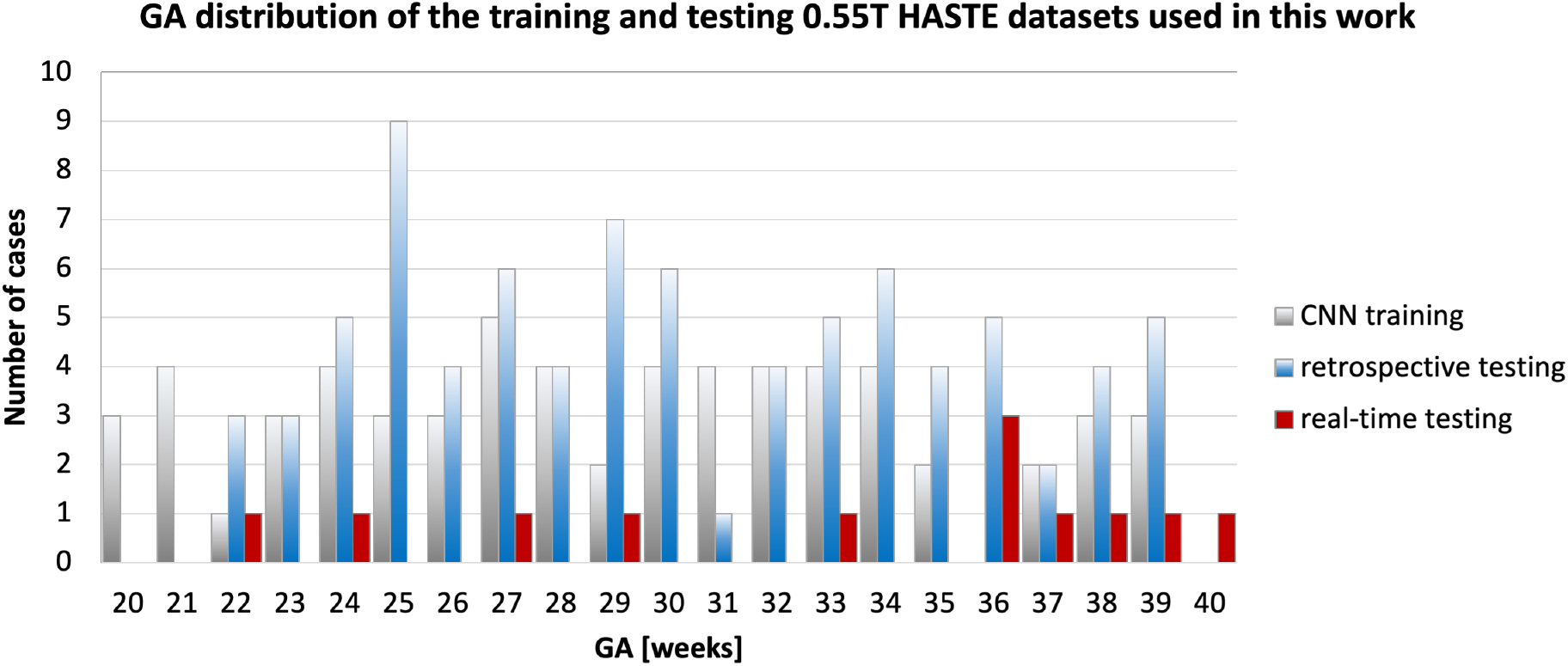
GA distributions of the 0.55T datasets used in this work for the training of the convolutional neural network (CNN), retrospective image quality evaluation, and real-time testing.

- the CNN training cohort: 384 stacks from 62 fetal datasets from 20-39 weeks GA range acquired during 05/2022-01/2023 period;
- the retrospective evaluation (quantitative and qualitative) cohort: 83 fetal datasets from 22-39 weeks GA range acquired during 02/2023-08/2023 period;
- the real-time testing cohort: 12 fetal datasets from 22-40 weeks GA range acquired during 01/2024-02/2024 period.

The main selection criteria for the retrospective testing cases were singleton pregnancy, > 22 weeks GA, no significant structural fetal pathologies, good in-plane image contrast with high signal-to-noise ratio (SNR), clear visibility of the whole fetus, and no breaks during ssTSE acquisition. This is a heterogeneous cohort with the maternal body mass index (BMI) varying between 22-43, different placental and fetal findings, and varying volume of amniotic fluid. Out of 88 originally inspected retrospective evaluation datasets, only 4 (5%) were excluded due to suboptimal image quality in the majority of stacks with severe in-plane signal loss caused by shading, extreme motion artifacts, or other acquisition-related factors. These cases were excluded because there was not enough image information (i.e., good-quality slices) for 3D SVR reconstructions. Furthermore, we did not include cases below 22 weeks due to the small size of the fetal organs compared to the intrinsic large slice spacing and resolution in 0.55T datasets that would be expected to compromise output reconstruction quality.

### 2.2 3D image-domain reconstruction

The proposed automated pipeline for combined 3D brain+body D/SVR reconstruction is summarised in Fig. 3. This is an extension of our previous work for automated 3D reconstruction of the fetal brain and thorax ^13,32^ with both brain and body regions of interest (ROIs) trained on 0.55T datasets using MONAI network implementations. It includes global 3D localisation of the brain and trunk in all stacks, followed by landmark-based reorientation to the standard radiological space, template selection, and classical rigid SVR and DSVR reconstruction. An additional reorientation step is applied to the final reconstructed images for refined alignment.

**FIGURE 3.**
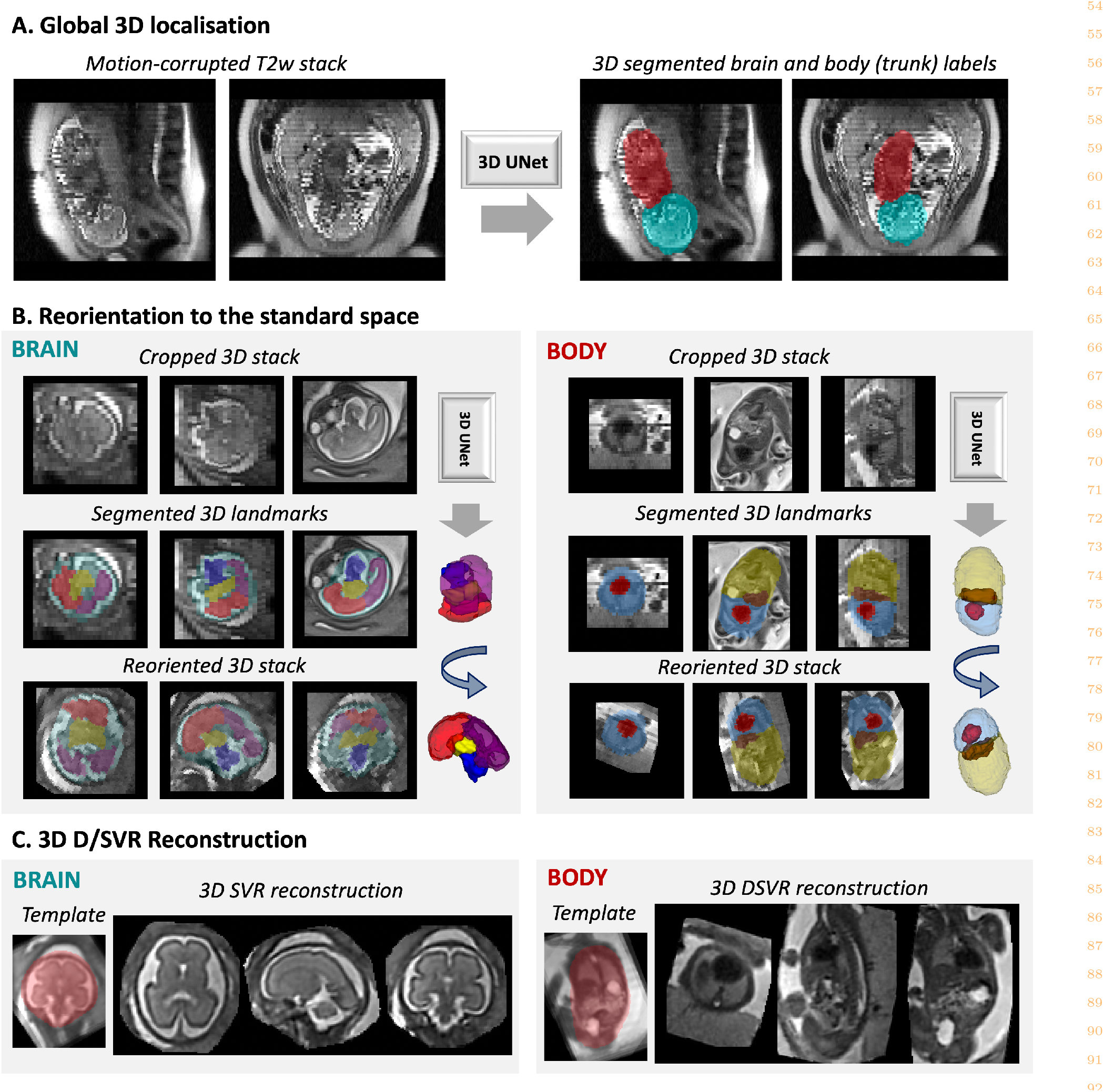
Proposed automated combined 3D brain+body D/SVR reconstruction pipeline for T2w 0.55T fetal MRI including: (A) global 3D localisation in motion-corrupted stacks; (B) 3D reorientation to the standard space; (C) average template creation and D/SVR reconstruction.

#### 2.2.1 Global localisation

Similarly to Uus et al. ^13^, the global localisation step is based on a 3D UNet ^35^ segmentation of brain and body (trunk) labels (Fig. 3 .A) that has already proven to be efficient for higher field strength fetal MRI. We used the classical MONAI ^34^ implementation and the network was trained on 384 stacks from 62 0.55T datasets with GA range 20-39 weeks, using standard augmentation (affine rotations, bias field, contrast adjustment, Gaussian noise) for 50 000 iterations. The labels for the training datasets were created semi-automatically based on manual refinement of the outputs of the existing network from ^13^.

#### 2.2.2 Reorientation to the standard space

Reorientation of the brain and body ROIs to the same canonical space of all stacks is an essential step to account for large rotation motion that cannot be corrected by classical registration. There have been many proposed alternative deep learning solutions for the reorientation of 2D fetal brain slices to the standard radiological space ^16,17,18^. In this work, we employ the existing pipeline for the fetal brain and thorax from our previous works ^13,32^ based on 3D landmarks for global 3D reorientation in raw stacks.

After global localisation, the stacks are cropped to the brain and body ROIs and passed to the 3D landmarks segmentation networks (Fig. 3. B). We selected 4 body landmarks (thorax, abdomen, heart, liver) and 4 brain landmarks (anterior brain, posterior brain, deep grey matter, cerebellum+brainstem). We used the classical 3D UNet implementations in MONAI for both networks. The body landmark network was trained on 173 0.55T cropped stacks with varying orientations and degrees of motion. The landmark labels for the training datasets were created manually. The brain landmark network was trained on 195 0.55T cropped stacks with varying orientations and degrees of motion. The labels were created using a semi-supervised approach as manually refined outputs from a separate in-house network pre-trained on fused Draw-EM labels from fetal dHCP data release ^‡^. Training included standard MONAI augmentation (affine rotations, bias field, contrast adjustment, Gaussian noise).

Next, the segmented landmarks are used for reorientation to the standard radiological space using classical point-based registration between centre points in the landmarks in stacks and atlases.

#### 2.2.3 3D D/SVR reconstruction

After reorientation, the ROI cropped and reoriented stacks are passed to the *stack-selection* ^13^ SVRTK function that performs additional rigid registration followed by template selection and rejection of outlier stacks based on a combination of normalized cross-correlation similarity and motion corruption metrics. The outputs are then used to create averaged 3D brain and body templates and the corresponding masks.

Next, the selected stacks and average ROI templates with masks are passed to the optimised rigid SVR ^8^ and DSVR ^9^ functions for 3D brain and body reconstructions (Fig. 3. C). The rigid SVR function reconstructs the large background ROI (in order to account for any mask imperfections) with the registration, bias correction, rejection of outliers, and intensity matching steps based on the brain mask ROI only. The optimised DSVR function is based on combined rigid+deformable slice-to-volume registration. Both DSVR and SVR reconstruction functions include an additional structure-based outlier rejection step ^9^. Taking into account the native stack resolution (1.2 × 1.2 × 4.5 mm), the output resolution for 3D reconstructed images was selected as 1.0 mm.

### 2.2.4 Implementation details

The deep learning modules were implemented based on MONAI^§^ framework. We used classical 3D UNet ^35^ architecture with five encoder-decoder blocks (output channels 32, 64, 128, 256, and 512), convolution and an upsampling kernel size of 3, ReLU activation, dropout ratio of 0.5, batch normalisation, and a batch size of 2. We employ an AdamW optimiser with a linearly decaying learning rate, initialised at 1x10-3, default beta parameters, and weight decay 1x10-5.

All pipeline components (deep learning and C++ reconstruction) are combined into one bash script. The code for the proposed brain and body reconstruction pipeline is publicly available at *auto-proc-svrtk* SVRTK GitHub repository^¶^.

The proposed 0.55T 3D D/SVR reconstruction pipeline is publicly available as a standalone docker application available at SVRTK docker repository^#^ that includes all required software installations as well as the network weights. The docker is fully CPU-based and is executed on the CPU for straightforward deployment purposes. The recommended minimum docker settings are > 20 GB RAM and 8 CPUs.

### 2.2.5 Evaluation metrics

The evaluation of the proposed D/SVR pipeline is based on a set of retrospective datasets. The quantitative evaluation is based on the localisation distance and reorientation rotation errors and label Dice. The quality scores for the output 3D D/SVR reconstructed images are: 1 - failed; 2 - poor, 3 - acceptable, 4 - good (similarly to the grading scheme in ^5^).

### 2.3 Gadgetron-based scanner D/SVR deployment

The entire online pipeline was implemented using Gadgetron. As in routine fetal MRI research and clinical protocols, T2w ssTSE data is acquired in 6 to 9 different orientations, planned according to maternal habitus and the fetal brain. As described above in Fig. 3 Step 1, the reconstructed stacks are exported to the dedicated GPU-equipped Gadgetron server immediately after acquisition, and D/SVR reconstruction is subsequently performed, triggered by a 5-second dummy sequence. The total reconstruction time is approximately 6:42±3:13 minutes, and the results are available immediately on the scanner console.

#### 2.3.1 Automated D/SVR activation

After all ssTSE stacks are acquired, a 5-second ‘launch’ sequence is run as part of the protocol (Fig. 3 Step 2). This establishes a connection between the scanner and the server and launches the SVR docker container on the server. An external-language interface Python Gadget was utilized to define this task and run it as a subprocess, allowing the acquisition and reconstruction of the subsequent sequences in the exam card. The acquired data from the launch sequence is discarded. The final reconstructions for the body and brain are written in NIfTI format to the Gadgetron server.

#### 2.3.2 Transfer of results to scanner

As a last step to bring the D/SVR reconstruction to the scanner host in a controlled way (ensuring that the reconstruction does not interfere with the scanning process), two fast ‘pull’ sequences are run towards the end of the protocol (24 seconds for pulling the brain volume and 1:03 minutes for the body volume). A modified, highly accelerated MP-RAGE sequence is employed for this purpose with the matrix size matched to the expected reconstruction. All brain reconstructions are resampled to an image matrix of 128x128x128 (128 slices) and the body reconstructions to 256x256x256 (256 slices) prior to this step - resampling facilitates the retrieval of the volumes in the scanner as having all brain/body reconstructed volumes with the same dimensions allows to keep the same field-of-view in the pulling sequences without compromising the quality of the results (e.g., cropped structures, missing slices). Similar to the sequence used to launch the docker, the purpose of these sequences is to establish the connection to the external server. In this connection, a Python Gadget was configured to disregard and overwrite the acquired data with the 3D SVR resulting volume matrices. Once the SVR volume is injected into the vendor image reconstruction chain and accessible by the scanner host, it is exported to the medical image database system alongside the acquired ssTSE images that were utilized to produce the reconstructed volume.

#### 2.3.3 Implementation details

The Gadgetron framework was installed on the dedicated external server and connected to the internal network of the MRI scanner. On the scanner side, the sequences were modified to link to the Gadgetron by inserting emitter and injector functors into the scanner processing pipeline. This allows the data to bypass specific sections of the pipeline by sending the raw data to the external server for processing (emitter functor) and receiving the image data (injector functor) with the integration of the image back into the scanner pipeline. Configuration files that specify crucial parameters such as the algorithms to be used during pipelines were created and stored in the scanner host to be accessed by the sequence. Additionally, the configuration files within the Gadgetron framework were developed for the real-time reconstruction and processing tasks and stored in the external server to be accessed when a connection to the server was established. For each of the three tasks presented in this work, a pair of configuration files was created - the configuration file stored in the scanner host, defined by the sequence, points to the reconstruction/processing configuration file that is stored in the external server. This file assembles the reconstruction and processing Gadgets the data streams through, including the Python-scripted external-language interface Gadgets that were developed for each task described in this work.

The code for the proposed Gadgetron-based D/SVR scanner integration for 0.55T fetal MRI is publicly available at *gadgetron-svrtk-integration* SVRTK GitHub repository^∥^.

## 3 RESULTS

The proposed Gadgetron-deployed brain+body D/SVR reconstruction pipeline was evaluated on 83 retrospective 0.55T datasets and with (*in utero*) prospective real-time testing of the integration step on 12 cases.

### 3.1 Retrospective testing

#### 3.1.1 Global localisation and reorientation

The results of the quantitative evaluation of the global brain and body localisation and reorientation on 30 datasets (with 60 stacks in total) within the 22-39 weeks GA range are presented in Tab. 1. The selected stacks have varying uterus and fetal acquisition planes. An evaluation was performed vs. manually created ground-truth 3D labels. Similarly to ^13^, the 3D UNet showed robust performance for both brain and body with relatively high Dice and average 6.067 ± 1.950 (brain) and 7.836 ± 3.027 (body) mm centre point distance errors for both early and late GA ranges. The proposed 3D localisation network ensures continuity of the individual labels in 3D space that does not require additional post-processing in comparison to the conventional 2D slice-wise fetal brain nnUNet approach ^15^. The quantitative evaluation of the reorientation of the brain and body to the standard radiological space was performed compared to the manually reoriented images. The landmark-based approach showed robust performance for both brain and body with 11.947 ± 6.626 (brain) and 15.549 ± 12.487 (body) degree rotation error ranges, which is an acceptable range for a classical registration method (Sec. 2.2.2). The higher rotation error for early GA datasets is due to the lower visibility of landmarks.

#### 3.1.2 3D D/SVR reconstruction

Evaluation of the automated D/SVR 0.55T reconstruction results was performed qualitatively on 83 datasets (not used in training) for both brain and body (AL, MH, AU) based on the quality scores defined in Sec. 2.2.5. The results in Fig. 4 .A-B demonstrate that the pipeline has relatively stable performance with acceptable and good reconstruction quality for both brain and body in the predominant (>85%) proportion of the datasets. The lower quality grades were in the early (due to small size) and late GA range (due to the lower cortical and tissue organ contrasts) and severe motion corruption cases with a high proportion of in-plane signal loss. Notably, the 0.55T reconstructed image quality is inherently lower (in terms of sharpness of features and tissue interfaces) vs. normal 1.5-3T results due to the expected difference in the original stack data quality.

**FIGURE 4.**
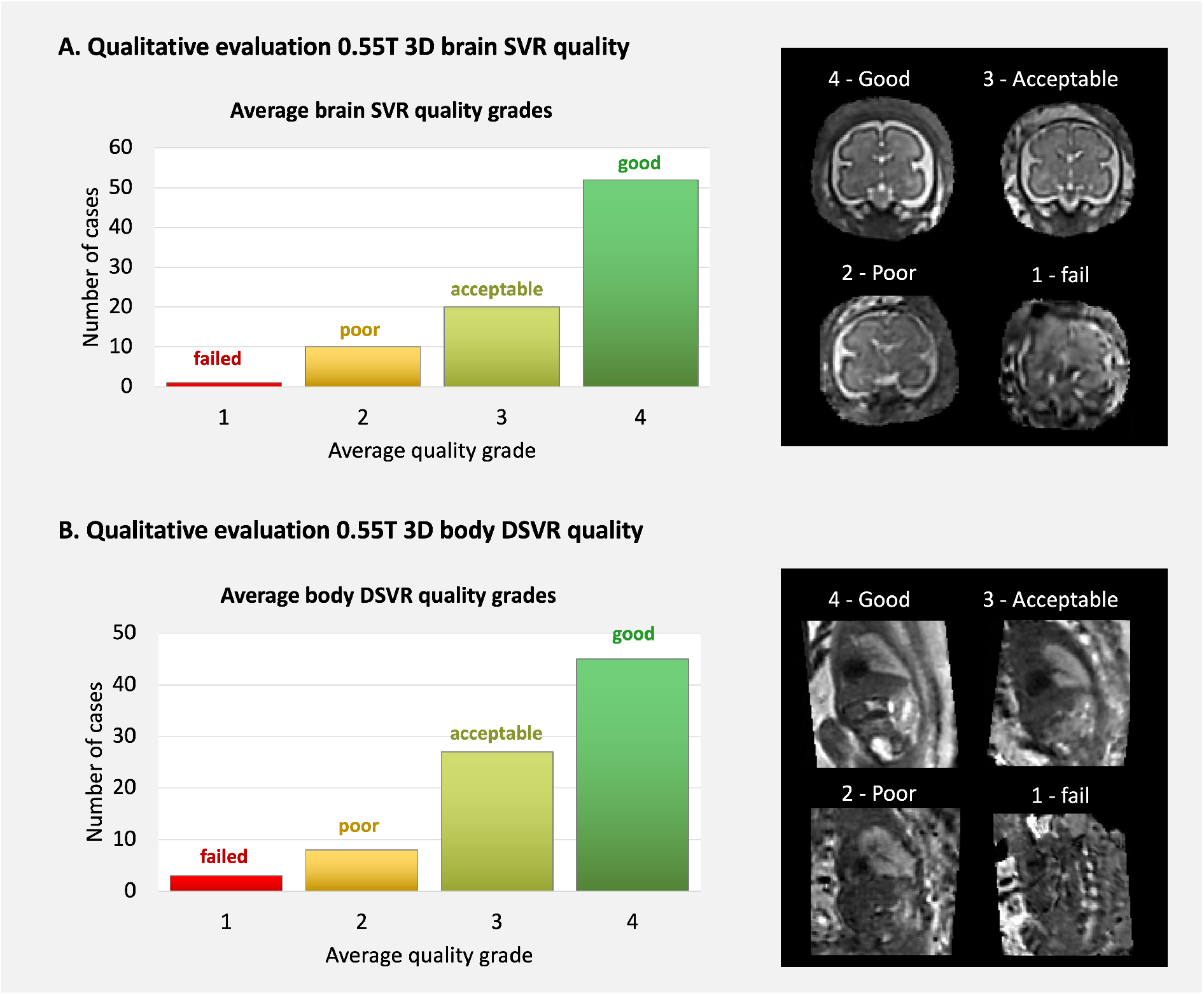
Retrospective qualitative evaluation of the brain (A) and body (B) reconstruction quality on 83 0.55T datasets from 22 - 39 GA range.

The main aims of this work are the general feasibility study for 0.55T and scanner-based integration of classical 3D reconstruction and do not include a detailed comparison with a large number of the recently proposed novel SVR approaches, i.e., other methods can be interchanged and deployed on the scanner in a similar format. The examples in Fig. 5 .A show a comparison between offline auto-SVRTK and the recent deep learning state-of-the-art GPU-based brain reconstruction NeSVoR method ^19^ on five 22-38 weeks GA cases, performed retrospectively. The outputs of both methods have similar global features but the tissue signal texture in SVRTK-reconstructed images is smoother but less noisy than in the NeSVoR results. This is potentially related to the differences in super-resolution reconstruction and regularisation methods as well as the lower SNR and resolution of 0.55T datasets. NeSVoR outputs cover of the brain ROI with a tight mask but SVRTK allows reconstruction of the background ROI. The average reconstruction time per case (with 9 stacks and 1 mm output resolution; default settings; on the same machine) was also within the similar range: 6-10 minutes (depending on GA) for auto-SVRTK and 9-10 minutes for NeSVoR (Fig. 5 .B).

**FIGURE 5.**
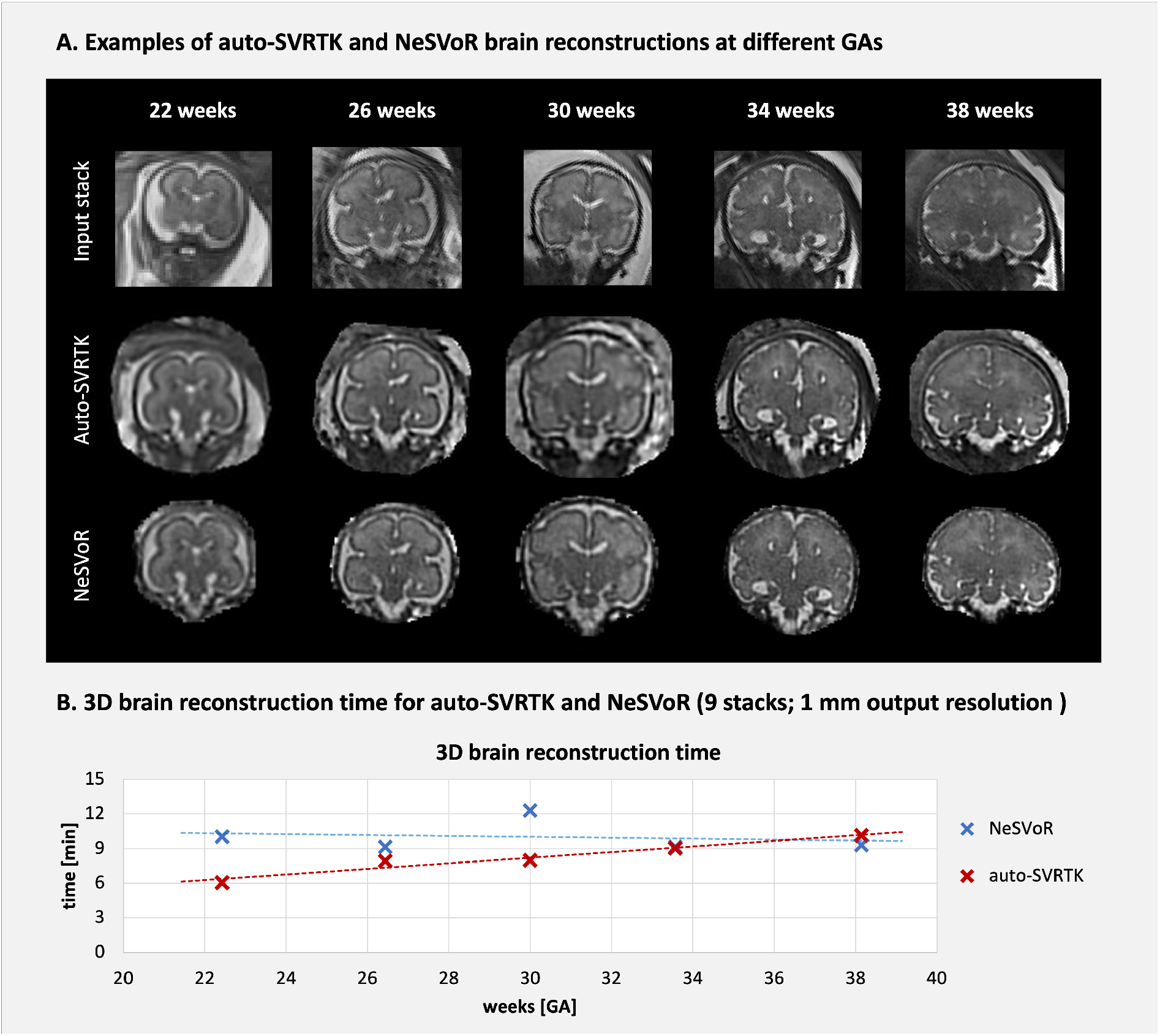
Examples of 3D auto-SVRTK vs. NeSVoR ^19^ brain reconstructions (A) and total reconstruction time vs. GA (B).

**TABLE 1.**
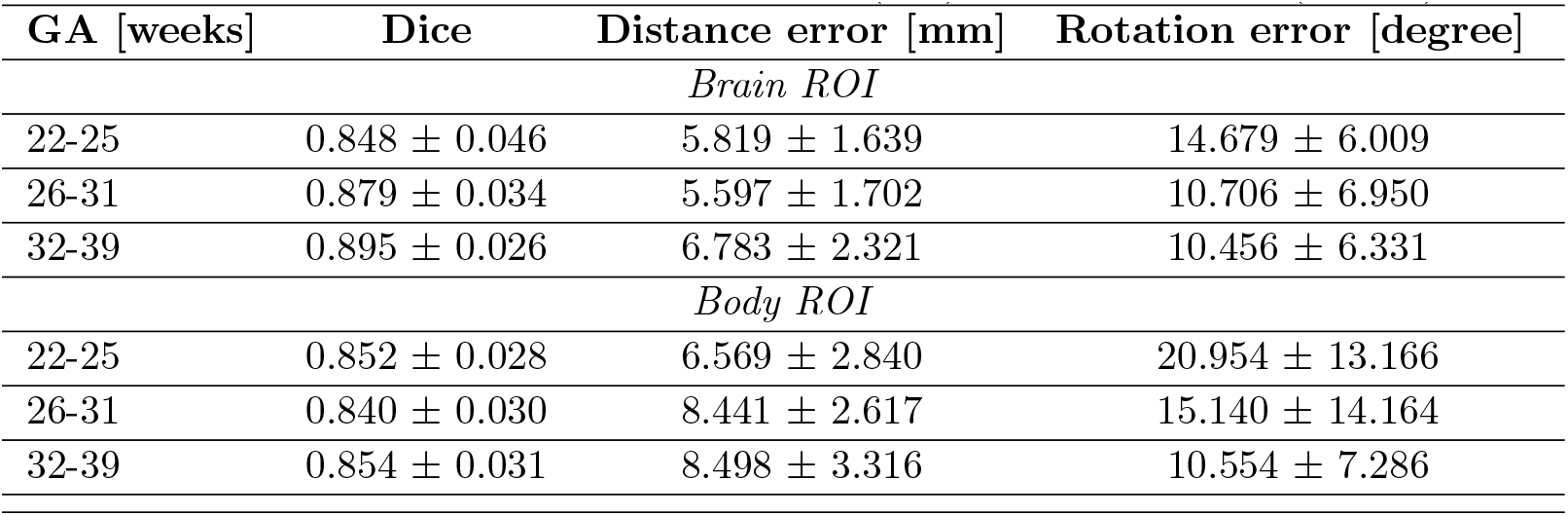
Retrospective quantitative evaluation of global localisation and reorientation to the standard space (B) on 30 datasets with 60 stacks in terms of Dice, localisation distance error (mm), and rotation error (degrees).

### 3.2 Gadgetron-based scanner D/SVR deployment

The 12 prospective cases all led to the motion-corrected super-resolved 3D reconstructions available on the scanner console during the fetal MRI scan, to view using all visualization options and to be archived with all acquired data at the end of the acquisition. The output image quality graded by 2 clinicians was within acceptable to good range. The launch and pull sequences on the protocol were moved freely by the radiographer performing the scan, with the launch sequence being acquired when a sufficient number of slices was obtained and the pull sequences performed after subsequent long diffusion sequences were acquired, at the end of the entire scan. Figure 6 A-C illustrates such a setup with the corresponding reconstructions for the brain (green boxes) and body (blue boxes). The time between the launch and the availability of the reconstruction for the brain was, on average, 7.23 minutes (median 6.39 min, std=3.03 min) and for the body 6.50 minutes (median 6.41 min, std=1.27 min), depending on the number of stacks, subject GA and computational system load. On average, no correlation of the processing time against BMI, GA or the number of stacks was observed (See Figure 6 D-F).

**FIGURE 6.**
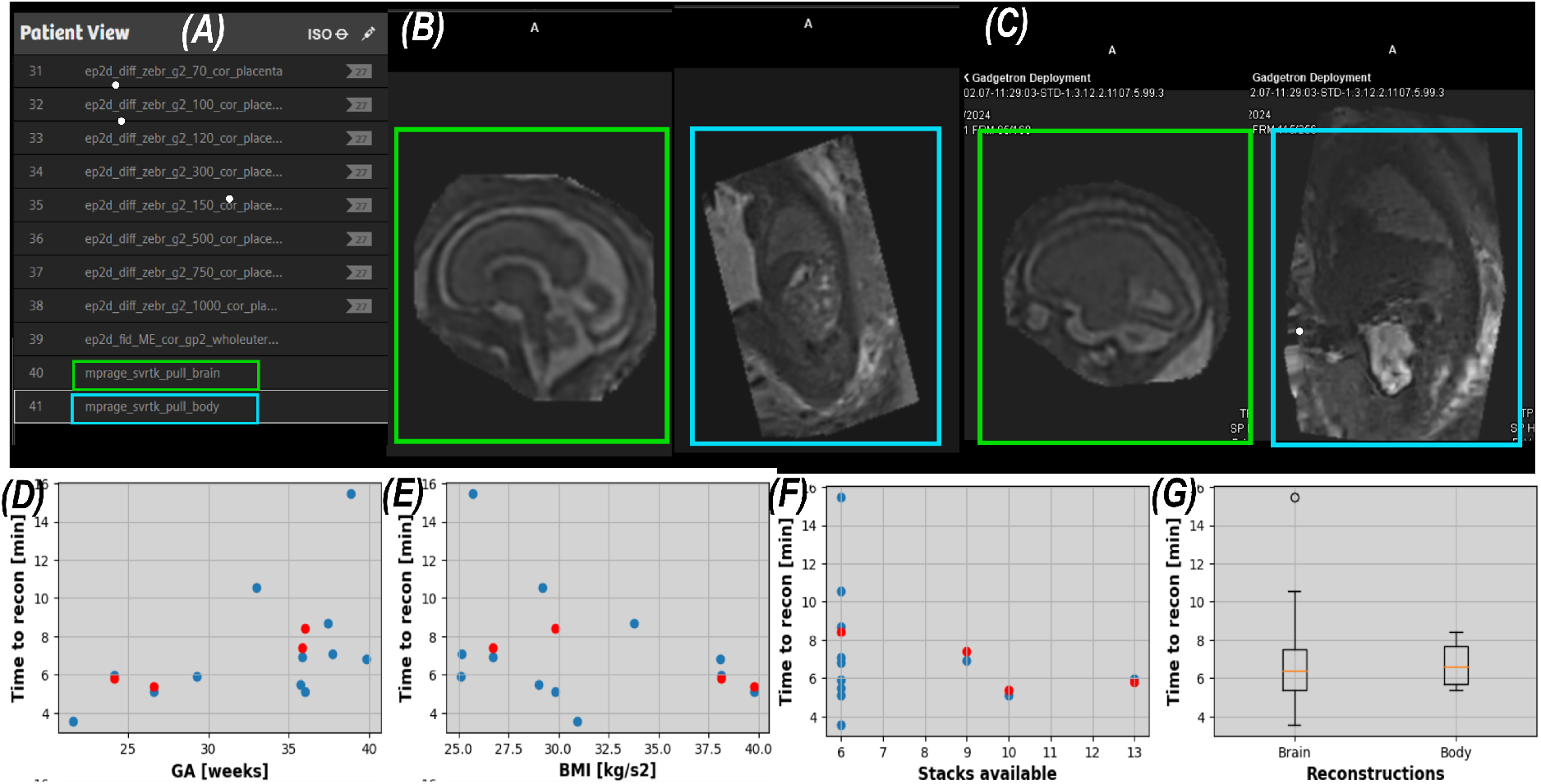
Results of the real-time deployment. (A) Screenshot of the end of the scanning protocol with the pull sequences. (B-C) Resulting brain (green box) and body (blue box) reconstructions on the scanner console during the MR scan for two cases. (D-F) Quantitative assessment of the time required for the online reconstructions against GA at scan, BMI and number of stacks. (G) Box plots illustrating the spread in time till the reconstruction was available online.

## 4 DISCUSSION

In this work, we designed and implemented the first pipeline for automated 3D D/SVR combined reconstruction of the fetal brain and body T2w at low-field MRI deployed on 0.55T scanner via Gadgetron resulting in motion-corrected 3D reconstructions being available on the scanner console during the examination. The main novel components are hence the compilation of fetal brain+body D/SVR methods into one combined pipeline, application of 3D image-domain reconstruction methods to low-field fetal MRI and integration into the scanner environment.

Quantitative and qualitative evaluation of the proposed automated D/SVR pipeline optimised for T2w 0.55T data on 83 retrospective cases demonstrated general suitability of using 3D reconstruction methods for low-field MRI. It also showed comparable performance with the most recent state-of-the-art deep leaning method for the fetal brain ^19^ in terms of both reconstruction quality and time. This suggests that the majority of the currently available SVR methods are generally interchangeable and that the SVR reconstruction quality is primarily defined by the input MRI data.

**TABLE 2.** Demographics and results for the prospective cases. Time refers to the time from the launch of the reconstruction on the scanner after the acquisition of the last ssTSE stack to the reconstruction being readily available on the scanner console computer. GA= Gestational Age, BMI= Body Mass Index, T1DM=Type 1 Diabetes Mellitus, IBD=Inflammatory Bowel Disease, CDH=Congential diaphragmatic hernia, PPROM = Preterm prolonged rupture of the membranes

|  | GA | BMI | Brain | Body | Stacks |
| --- | --- | --- | --- | --- | --- |
|  | [weeks] | [kg/m<sup>2</sup>] | [min] | [min] |  |
| 1-healthy | 37.71 | 25.17 | 07:05 | - | 6 |
| 2-healthy | 37.42 | 33.80 | 08:42 | - | 6 |
| 3-healthy | 35.7 | 29.0 | 05:29 | - | 6 |
| 4-T1DM | 21.57 | 30.92 | 03:37 | - | 6 |
| 5-healthy | 33 | 29.2 | 10:36 | - | 6 |
| 6-IBD | 29.3 | 25.1 | 04:52 | - | 6 |
| 7-healthy | 39.85 | 38.09 | 06:48 | - | 6 |
| 8-healthy | 38.85 | 25.71 | 15:28 | - | 6 |
| 9-Fibroids | 35.85 | 26.73 | 06:54 | 07:26 | 9 |
| 10-CDH | 26.57 | 39.79 | 05:07 | 05:25 | 10 |
| 11-healthy | 36 | 29.83 | 05:01 | 08:35 | 6 |
| 12-PPROM | 24.14 | 38.16 | 05:55 | 05:56 | 13 |

This prototype automated pipeline also allowed integration of D/SVR methods directly into the scanner environment enabling 3D reconstructions to be visualised on the scanner console during the examination. The results of real-time *in-utero* prospective testing on 12 cases confirmed the robustness of the method, no interference with the scan acquisition and the ability to achieve good data quality. The 3D brain reconstruction from 6 stacks was available on the console within 7 minutes on average, and only one case took > 10 minutes, which was related to a general slowed-down console computer on this day.

### Limitations and future work

Since the primary aims of this work are the general D/SVR feasibility assessment for 0.55T and real-time integration we did not include an assessment of the pipeline performance on the extreme motion and signal loss datasets. In clinical practice, these cases normally represent only a small proportion of datasets and should potentially be addressed by real-time acquisition quality control since the lack of original 2D slice image information will make any 3D reconstructed images unreliable by definition.

Our future work will also focus on detailed technical formalisation of the minimum input image requirements and an automated real-time stack quality classification module that could also provide active guidance for the re-acquisition of motion corrupted stacks ^22^.

One of the potential limitations of this work is the reliance on the classical D/SVR methods for the reconstruction part and the expected sensitivity of the landmark reorientation method to low-quality stacks. Even though the full pipeline is operational, integration of more advanced deep learning super-resolution reconstruction and pose estimation methods ^19^ would be beneficial for robust performance for severe motion corruption cases. This will be addressed in our future work based on a thorough investigation of the optimal D/SVR reconstruction pipeline modules for structural MRI along with the extension with automated organ parcellation ^36^ for volumetry reporting during scanning. Furthermore, we will explore solutions for deep learning image recovery specific for datasets with severe signal and contrast loss.

The fact that the proposed structure of the Gadgetron-based scanner deployment relies on the external launch of the D/SVR docker application can also be considered a form of limitation since the activation and the transfer of results back to the scanner rely on the file availability logic. One of the possible solutions could be based on more advanced integration of the reconstruction code into the Gadgetron interface: e.g. dynamic masking, reorientation ^21^ and sequential addition of stacks to the reconstruction function.

While implemented here for a 0.55T system from one vendor, generalisation of the pipeline for other systems operating at 1.5-3T field strengths from a wider range of vendors is a logical next step, supported by the use of open-source tools at every step other than the MR sequence itself.

Another essential aspect is the qualitative assessment of 3D D/SVR images at 0.55T in terms of added diagnostic value and image quality limitations. This will require a wider clinical analysis of brain and body reconstructions along with a comparison to 1.5-3T data.

## 5 CONCLUSIONS

This study shows the feasibility of a real-time scanner-integrated solution for 0.55T fetal brain and body 3D reconstruction. The initial step towards integration of 3D D/SVR reconstruction methods directly into the clinical environment has been developed and has the potential to enable straightforward processing and reporting of fetal MRI scans. Our future work will focus on further optimisation and integration of the D/SVR reconstruction and image quality control into the scanner environment for real-time processing.

## Data Availability

The code for the proposed Gadgetron-based D/SVR
scanner integration for 0.55T fetal MRI is publicly
available at gadgetron-svrtk-integration SVRTK GitHub
repository.

https://github.com/SVRTK/gadgetron-svrtk-integration

## Abbreviations

MRI: magnetic resonance imaging
SVR: slice-to-volume registration

## ACKNOWLEDGMENTS

We thank everyone who was involved in the acquisition and analysis of the datasets at the Department of Perinatal Imaging and Health at Kings College London and St Thomas’ Hospital. We thank all participants and their families.

The authors acknowledge the invaluable help of the radiographers and midwives while acquiring the data presented here.

The views expressed are those of the authors and not necessarily those of the NHS, the NIHR or the Department of Health.

## Author contributions

AU and SNS contributed equally to this work and prepared the manuscript. AU designed and implemented the automated reconstruction pipeline and performed the retrospective evaluation. SNS designed and implemented the Gadgetron scanner integration pipeline. JAV designed the acquisition protocols and contributed to scanner integration. KP, MH and AL contributed to the acquisition and analysis of the results. MM and KC contributed to the acquisition of the datasets. TR contributed to the development of the automated reconstruction pipeline. HS contributed to SVR testing. SM contributed to the development of acquisition protocols and scanner integration. ZN contributed to the Gadgetron pipeline. MD, JVH, MR, and LS supervised various components of the project. JH designed the acquisition protocols and scanner integration pipeline, provided datasets, prepared the manuscript and supervised the project.

## Financial disclosure

None reported.

## Conflict of interest

The authors declare no potential conflict of interests.

## ORCID

*Alena U. Uus*\*

*Sara Neves Silva*\*

*Jordina Aviles Verdera*

*Kelly Payette*

*Megan Hall*

*Kathleen Colford*

*Aysha Luis*

*Helena S. Sousa*

*Zihan Ning*

*Thomas Roberts*

*Sarah McElroy*

*Maria Deprez*

*Joseph V. Hajnal*

*Mary A. Rutherford*

*Lisa Story*

*Jana Hutter*

## Footnotes

* VRTK toolbox repository: https://github.com/SVRTK/SVRTK

† Gadgetron repository: https://github.com/gadgetron/gadgetron

‡ dHCP fetal MRI data release: https://www.developingconnectome.org/project/

§ MONAI framework repository: https://github.com/Project-MONAI/MONAI

¶ SVRTK auto-proc-svrtk repository: https://github.com/SVRTK/auto-proc-svrtk

\# SVRTK auto D/SVR reconstruction docker: https://hub.docker.com/r/fetalsvrtk/svrtk; tag general auto amd)

∥ SVRTK gadgetron-svrtk-integration repository: https://github.com/SVRTK/gadgetron-svrtk-integration

## Notes

**Funding Information** This work was supported by the Wellcome Trust, Sir Henry Wellcome Fellowship to JH [201374/Z/16/Z], the UKRI FLF to JH [MR/T018119/1], DFG Heisenberg [502024488] the High Tech Agenda Bavaria to JH, the NIHR Advanced Fellow-ship to LS [NIHR3016640], the MRC grants [MR/W019469/1] and [MR/X010007/1], and the Wellcome/EPSRC Centre [WT203148/Z/16/Z].

### Competing Interest Statement

Sarah McElroy - Siemens Healthineers

### Funding Statement

This work was supported by the Wellcome Trust, Sir Henry Wellcome Fellowship to JH [201374/Z/16/Z], the UKRI FLF to JH [MR/T018119/1], DFG Heisenberg [502024488] the High Tech Agenda Bavaria to JH, the NIHR Advanced Fellowship to LS [NIHR3016640], the
MRC grants [MR/W019469/1] and [MR/X010007/1], and
the Wellcome/EPSRC Centre [WT203148/Z/16/Z].

### Author Declarations

The fetal MRI data used in this study were acquired at St.Thomas' Hospital, London as part of the ethically approved MEERKAT [REC: 21/LO/0742], MiBirth [REC: 23/LO/0685] and NANO [REC: 22/YH/0210] studies.

